# “Comparative analyses revealed reduced spread of COVID-19 in malaria endemic countries”

**DOI:** 10.1101/2020.05.11.20097923

**Authors:** Azhar Muneer, Kiran Kumari, Manish Tripathi, Rupesh Srivastava, Asif Mohmmed, Sumit Rathore

## Abstract

In late 2019, SARS-CoV-2 (Severe Acute Respiratory Syndrome Coronavirus 2) infection started in Hubei province of China and now it has spread like a wildfire in almost all parts of the world except some. WHO named the disease caused by SARS-CoV-2 as COVID-19 (**Co**rona**Vi**rus **D**isease-20**19**). It is very intriguing to see a mild trend of infection in some countries which could be attributed to mitigation efforts, lockdown strategies, health infrastructure, demographics and cultural habits. However, the lower rate of infection and death rates in mostly developing countries, which are not placed at higher levels in terms of healthcare facilities, is a very surprising observation. To address this issue, we hypothesize that this lower rate of infection is majorly been observed in countries which have a higher transmission/prevalence of protozoan parasite borne disease, malaria. We compared the COVID-19 spread and malaria endemicity of 108 countries which have shown at least 200 cases of COVID-19 till 18^th^ April 2020. We found that the number of COVID-19 cases per million population correlates negatively with the malaria endemicity of respective countries. The malaria free countries not only have higher density of COVID-19 infections but also the higher case fatality rates as compared to highly malaria endemic countries. We also postulate that this phenomenon is due to natural immune response against malaria infection, which is providing a heterologous protection against the virus. Unfortunately, there is no licensed vaccine against SARS-CoV-2 yet, but this information will be helpful in design of future strategies against fast spreading COVID-19 disease.

## Introduction

SARS-CoV-2 (Severe **A**cute **R**espiratory **S**yndrome **Co**ronavirus **2**) infection is affecting almost all countries in the world. After being reported by WHO in early January 2020 about human to human transmission of the virus, till now the spread is on increase day by day (www.who.int). WHO named the disease caused by SARS-CoV-2 as COVID-19 (**Co**rona**Vi**rus **D**isease-20**19**) and declared it a global pandemic in February 2020. However, there are striking differences in the behavior of COVID-19 spread in different countries of different continents. For example, United States of America is been shown to have highest number of cases and the spread is pretty fast with high mortality rates (www.cdc.gov). On the other hand South Korea and Taiwan, despite having initial cases have been reported too early, show a slower spread of infection and less mortality rate. Medical infrastructure and correct timing of restrictions on public gatherings and movement can be listed as factors which has led to such differences in the menace created by this virus [1].

We propose an alternate hypothesis regarding the spread of COVID-19, which is that there is limited spread of the disease in malaria endemic countries as compared to malaria free countries. Malaria is a vector borne disease caused by apicomplexan parasite *i.e. Plasmodium* sp. Malaria is a deadly disease that causes about 228 million cases and more than 405,000 deaths per year globally as per WHO report 2019. Malaria distribution across the world can be divided as endemic regions and non-endemic regions as per the number of cases reported per million of the population (WHO malaria report 2019). In endemic areas, malaria is being reported to occur repeatedly throughout the year and hence the population in these areas get infection at regular intervals [2]. Malaria cases have also been governed by presence of vector, Anopheles mosquitoes in that region as they are the prime carriers of the parasite. Some countries are free of malaria due to the climate conditions, lack of vector and successful malaria eradication efforts which include North America and Europe along with others [3]. Population in malaria endemic countries are been reported to achieve a natural protection against future infection by generating combination of Th1 and Th2 response against the pathogen. This anti-malaria immunity can stay for longer durations, upto 3 years in some cases [4]. Lower number of COVID-19 cases are being reported from Malaria endemic countries (WHO COVID-19 status report). In the light of the fact that immunity against one pathogen can be effective in controlling infection by another pathogen, as also suggested by various studies including BCG vaccination [5], [6]; we hypothesize that anti-malaria immunity may play a role in slowing down the spread of COVID-19 viral infection.

In order to understand the correlation between less numbers of covid-19 cases in malaria endemic regions, we carried out this study by analyzing current epidemiological data of the two diseases and tried to identify the possible mechanisms behind such observation. The study will be helpful in understanding the natural resistance against the COVID-19 infection and can provide useful information regarding better management of this pandemic.

## Material and Methods

We collected the data about Malaria infection across the globe from WHO report 2019. The data for COVID-19 infection is been obtained from worldometer (https://www.worldometers.info/coronavirus/) as of 18^th^ April 2020. Data was plotted and analyzed using Graphpad prism and R package based statistical analyses.

## Results

### Distribution of malaria and COVID-19 shows drastic contrast

We plotted the COVID-19 trend and malaria burden on world maps and a straight look at them highlights the stark contrast between trends for the two diseases. The less spread of COVID-19 is seen in third world countries including Africa, Asia and Latin America (Figure 1) which are clearly seen having greater burden of malaria. Whereas, in European countries and USA, the COVID-19 spread is very high while these countries/regions are almost malaria free. It points towards possible correlation between less infectivity of COVID-19 and high rates of malaria.

**Figure 1:**
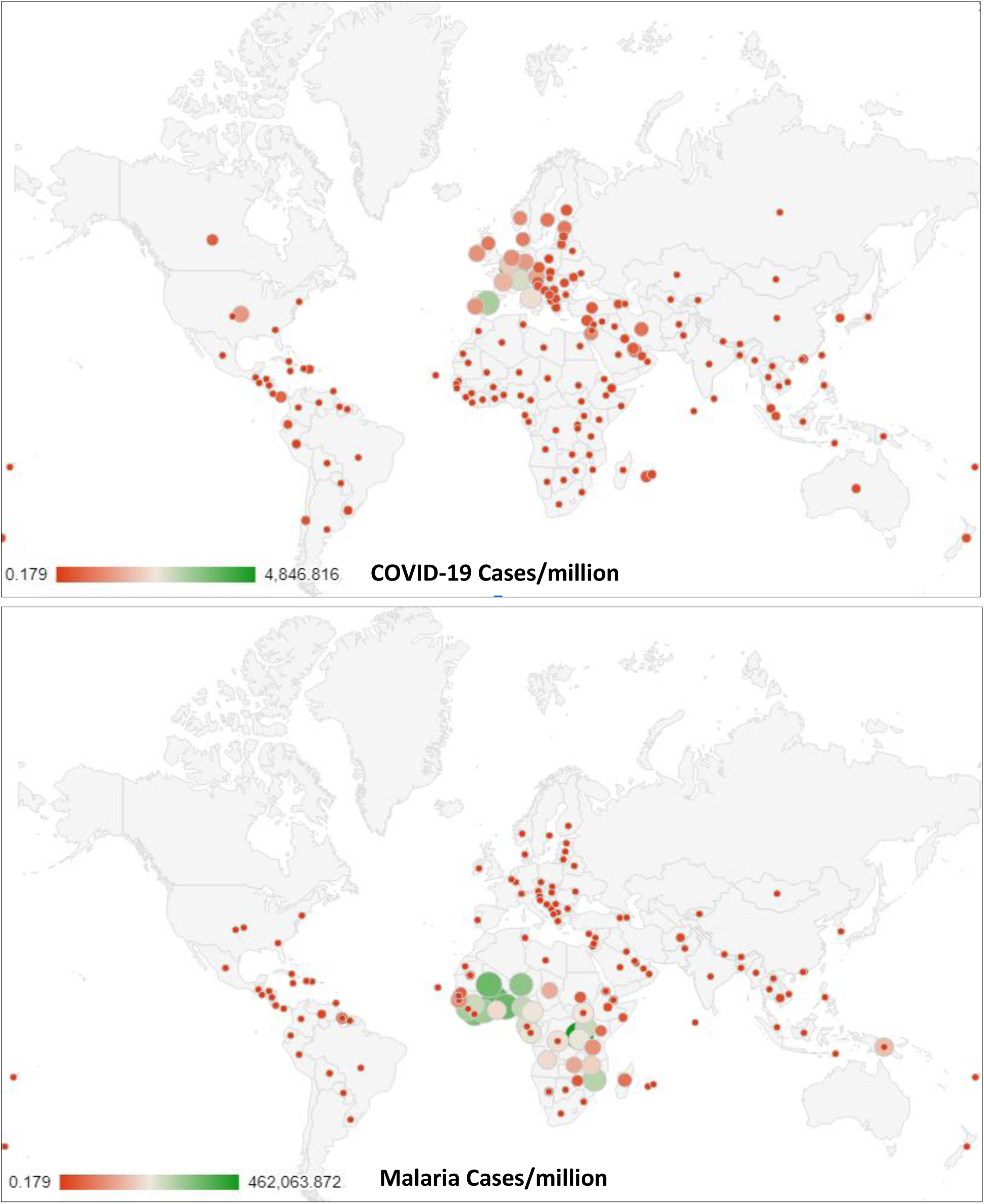
World maps displaying Number of cases worldwide (a) COVID-19 cases/million, (b) malaria cases/million as per WHO report 2018.

### COVID-19 burden is negatively correlated with malaria burden

In order to get a quantitative understanding of correlation between the burdens of COVID-19 and malaria, we compared the COVID-19 cases/million populations till 18^th^ April 2020 with malaria cases/million (in 2018) of 108 countries reporting at least 200 COVID-19 cases and having a population of at least 0.5 million. We have excluded countries which have not reported at least 200 of COVID-19 cases as those countries majorly have imported cases of COVID-19 rather than community spread, and/or also due to underreporting by the local administration. Also, for studying correlation between malaria burden and COVID-19 burden, it is necessary to not consider countries with only imported cases or very few local infections. By statistical analyses of data from these 108 countries we found that there is an inverse correlation between incidences of malaria in 2018 and COVID-19 infections (r= - 0.15, p= 0.02). Further we grouped these 108 countries into four categories based on extent of malaria burden; countries with, (i) no malaria, (ii) 1 to 1000, (iii) 1000 to 100 thousand and (iv) >100 thousand malaria cases/million in 2018, each category having 76, 14, 10 and 8 countries respectively and calculated the combined COVID-19 burden of each category. The total population of countries in each of these categories are 3400, 768, 2255 and 426 million respectively (Figure 2A). As shown in figure 2B, the countries having no malaria burden at all have an average COVID-19 burden of 576.6 cases/million whereas countries with malaria burden 1 to 1000, 1000 to 100 thousand and >100 thousand cases/million have average COVID-19 burden at 67.4, 31 and 10.2 cases/million respectively. Further, the shares of each category in total number of COVID-19 cases are 94%, 2.5%, 3.3% and 0.2% for groups, respectively. These data clearly shows a negative correlation between malaria endemicity and COVID-19 burden, suggesting the possibility of role of anti-malaria immunity in curtailing the SARS-CoV-2 spread.

**Figure 2:**
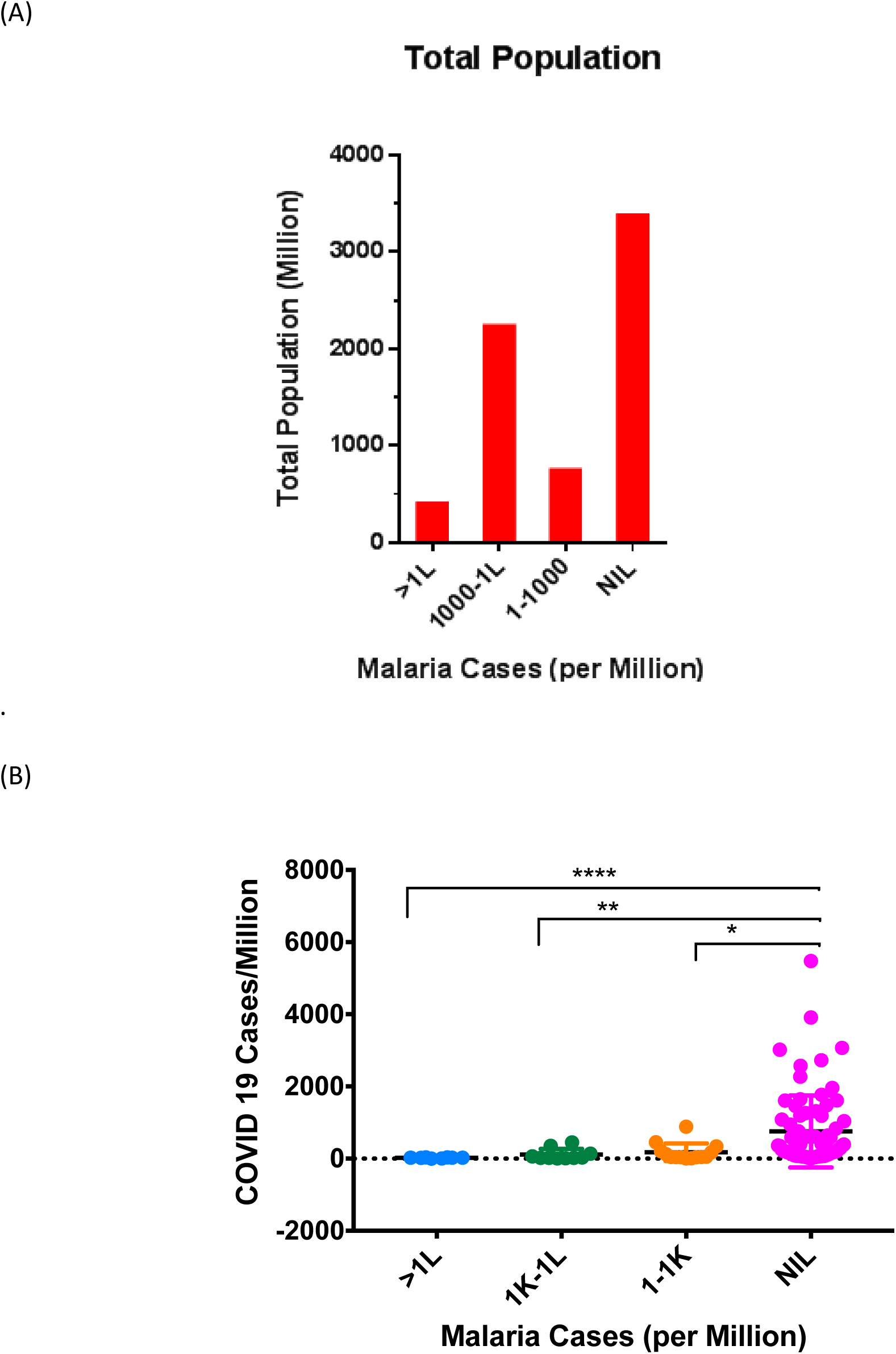
(A) Bar Graph representing total population of group of countries having different number of malaria cases as per WHO report 2018 (B) Scatter plot representing Number of COVID-19 cases/million (>200 cases) in group of countries having different number of malaria cases.

### Malaria free countries have high prevalence of COVID-19 Infections

Next, in order to have more detailed understanding of the correlation between malaria endemicity and COVID-19 cases, we did a country-wise analysis of COVID-19 and malaria burdens in 108 countries selected above. Out of 108 countries, total 76 countries are malaria free countries whereas remaining 32 countries are mild to highly endemic for malaria. We analyzed number of COVID-19 cases in 76 malaria free countries (as per WHO Malaria report 2018) and observed that most of the European countries have reported more than or equal to 1000 cases/million population (Figure 3). Luxembourg has the highest density of COVID-19 cases at >5000 cases/million and is followed by Spain, Switzerland and Belgium each showing >3000 cases/million. Italy, Ireland and France each reported >2000 COVID-19 cases/million. United States of America has maximum number of COVID-19 cases in the world but owing to its large population, its COVID-19 burden is 1958 cases/million (Figure 3A). A total 37 of these countries have shown COVID-19 burden between 100 to 1000 cases/million with majority of countries from malaria free Middle East Asia falling in this category. Remaining 19 malaria free countries have shown <100 cases/million and include countries like Lebanon, Kyrgyzstan, Ukraine, Tunisia, Japan, Iraq, China and Sri Lanka etc. Countries form middle east (i.e. Lebnanon, Egypt, Jordan and Iraq) and African regions (i.e. Tunisia) have lesser number of infections despite being malaria free regions. These less number of COVID-19 cases in these regions can be attributed to lot of factors, including crippled medical infrastructure due to war and less number of tests being conducted and lack of proper reporting by their administrations. However, these countries still have reported roughly 40-60 COVID-19 cases/million of the population. China is surprisingly seen among this group having lower COVID-19 burden at 57.43 cases/million, which could be because of many reasons. Firstly, the SARS-CoV-2 infection in humans originated in Wuhan city of China’s Hubei province, so it was very easy for Chinese administration to contain the disease in local area and directing the mitigation efforts to one particular area rather than at scattered hotspots as in case of other countries. In addition to this, the vast landscape of China along with largest population in the world (>1.4 billion) has brought down COVID-19 cases/million despite having ~82,000 cases of COVID-19 placed at 7^th^ position in the world in terms of total number of cases. Sri Lanka is another outlier in this group having only 11.6 COVID-19 cases/million which could be explained by the fact that it has become malaria-free just 3-4 years back and had significant malaria burden earlier, along with this, the local government’s well-articulated and timely mitigation efforts further helped in halting the SARS-CoV-2 spread.

**Figure 3:**
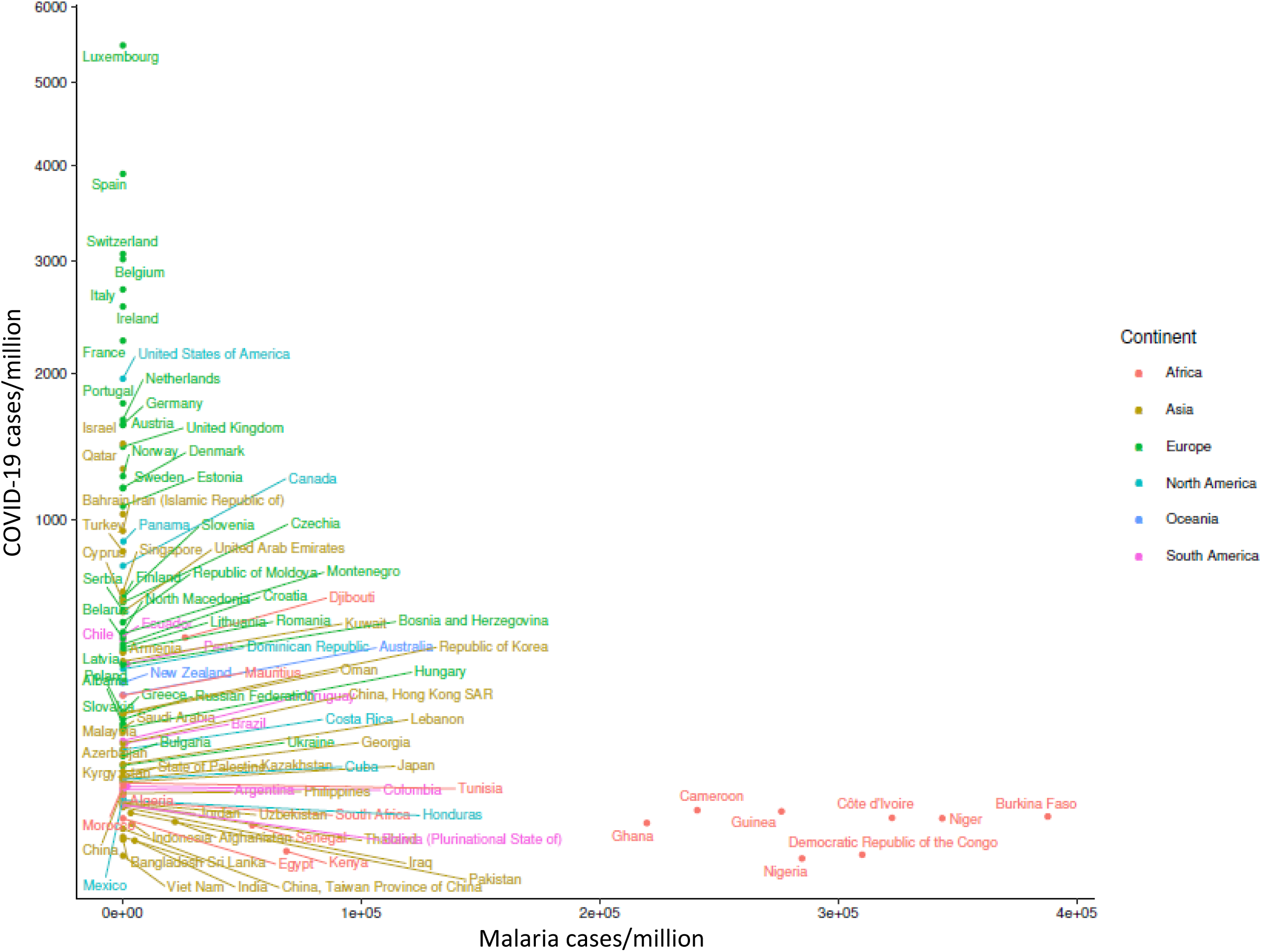
Bubble plot representing number of COVID-19 cases and malaria infections among 108 countries. Size of the bubble represent the number of cases reported from that respective country. Colour of bubbles represent different continents.

**Figure 4:**
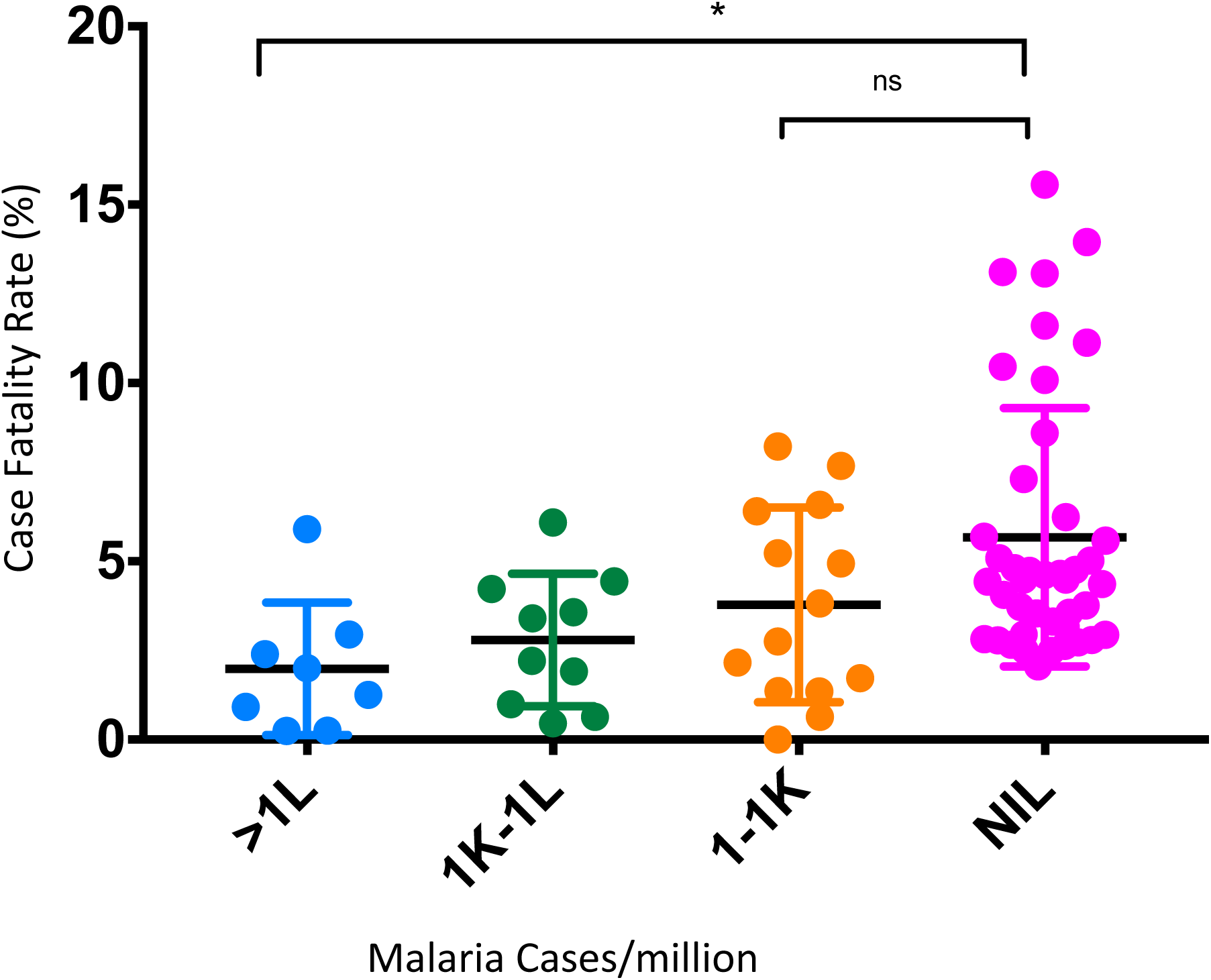
Scatter plot representing Case fatality rate (deaths/cases) for different group of countries as per there malaria burdens.

### Malaria endemic countries have reduced prevalence of COVID-19

Similarly, we analyzed COVID-19 cases/million and malaria cases/million for 32 countries having malaria (Figure 3). Countries with malaria burden of 1-1000 cases/million showed a mixed kind of response to COVID-19 with burden ranging from 2.8 and 9.6 cases/million for Vietnam and Bangladesh respectively, to 452 and 883 cases/million in Ecuador and Panama respectively (Figure 3). High density of COVID-19 cases in Ecuador and Panama could be due to significantly lower malaria burden (95.1 and 185 cases/million, respectively). Among these, Republic of Korea has the highest total number of COVID-19 cases (10,613 cases) but its well praised mitigation efforts and medical interventions have helped it to significantly curtail the spread of SARS-COV-2. Also, it has a negligible malaria burden of only 9.8 cases/million. Countries with yearly malaria infections between 1,000 to 100 thousand cases/million showed a very low numbers of COVID-19 burden ranging from ~4.3 to 446.8 cases/million. Out of these 10 countries, Djibouti, Peru and Brazil have highest density of COVID-19 at nearly 447, 352 and 137 cases/million respectively. Djibouti surprisingly has very high COVID-19 cases/million despite having high malaria burden of ~26,000 cases/million but it could be explained by looking at the number of actual COVID-19 cases and its population. It has reported only 435 cases of COVID-19 but due to its smaller population of only ~973,000, the COVID-19 burden expressed as cases per million, become high enough to create delusion. Peru on the other hand despite having malaria burden of 1798 cases/million showed higher COVID-19 burden at 352 cases/million which could be attributed to its demography and lack of good healthcare infrastructure culminating into inefficient screening and reporting. Brazil on the other hand showed COVID-19 burden of 137 cases/million which is on higher side in the group despite moderate malaria burden and could be attributed to the lack of political will in containing the spread of SARS-COV-2. Remaining seven countries of this group have negligible COVID-19 burden of ≤61 cases/million which correlated well with high burden of malaria in these countries. Further, eight countries which are highly endemic for malaria having malaria burden more than 100 thousand cases/million showed a minimal burden of COVID-19 ranging from 2 to 31.6 cases/million. All of these countries showed less than 1000 total cases of COVID-19 thereby, suggesting the negligible or no spread of this disease among its population (figure 3). Overall, these quantitative analyses support our hypothesis that extent of malaria burden does play a significant role in curtailing the spread of COVID-19 across the world.

### Malaria free countries have higher COVID-19 case fatality rate (CFR)

Relatively lower numbers of reported COVID-19 infections in a county, is also been attributed to less number of tests being conducted, as the case for several Asian and African Countries. In such situations, case fatality rate (CFR) will be a better assessment tool to understand the fatality of COVID-19 infection in malaria endemic as well as malaria free countries. For this, we analyzed the case fatality rate (expressed as percentage of COVID-19 positive cases who succumbed to the disease) for the four groups of countries divided on the basis of malaria burden as described in the previous sections. In total population of malaria free seventy six countries the COVID-19 case fatality rate is 6.63% and is highest among all the groups (Figure 4). In the second group having mild malaria burden of 1 to 1000 cases/million, the COVID-19 case fatality rate of 3.88% is observed. Similarly, for the group with moderate malaria burden of 1000 to 100 thousand cases/million, the COVID-19 case fatality rate of 4.63% is observed. Whereas, in the group having severe burden of malaria >100 thousand cases/million, the COVID-19 case fatality rate is just 2.58% which is the lowest among four groups and also significantly lower than that of malaria free countries (Figure 4). These case fatality statistics further support that along with lesser spread, the fatality of SARS-CoV-2 is also mild in populations having higher malaria burden.

## Discussion

COVID-19 infection is spreading in the world very rapidly and had claimed about 0.15 million human lives till 18^th^ April 2020. Based on our analyses we hypothesize that there is natural resistance against the spread of this viral disease in malaria endemic countries, which can be partially explained by immunity generated against recurrent malaria infections in these endemic regions [7, 8]. The countries of African and Asian regions where malaria is prevalent, have shown considerably fewer cases of COVID-19 infections as well as mortalities. Whereas, majority of the European countries, which haven’t seen much of malaria infection due to *Plasmodium*, have reported much higher number of COVID-19 cases and associated mortalities. Hence, the population that has always remained unexposed to malaria antigens thereby lacking anti-malarial immunity, showed a significantly higher burden of COVID-19 in comparison to populations which are exposed to malaria infections. It points toward resistance being provided by antimalarial immunity against the spread of COVID-19 infection.

Cross immunity against infections is a phenomenon which is been shown in the past [6], and it’s a well established fact now that one pathogen can generate immunity which can be deleterious for another pathogen [6]. In addition, it is being shown that vaccine against one pathogen can provide a long lasting immunity against another pathogen [5]. In this antagonistic phenomenon, pathogen of certain species can inhibit the infection by another one through inhibiting the invasion, survival and/or reproduction of another pathogen [9]. Different parasites are been reported to reduce the severity of viral infections, one of such example comes from Giardia which can reduce the severity and infection of rotavirus if concurrent infection takes place [10]. *Plasmodium* infection also can influence the infection caused by Chiukungnya virus [11] where the protection is attributed to the inhibition of CD8+ cell migration due to *Plasmodium* infection [11].

SARS-CoV has evolved a way to evade innate antiviral type I interferon responses of host cells in order to prolong viral replication and survival [12]. Recent studies have also suggested that dysregulated type I and II interferon responses may culminate in to failure of the switch from hyperinnate immunity to protective adaptive immune responses in SARS patients with a poor outcome [13] It is only in convalescent SARS-COV-2 patients that increased levels of IFNγ, IL-4 and IL-10 were observed [14] [15] [16]. It points out to host response towards SARS-COV-2 infections and protective role of IFN-γ and IL-10 against the SARS-COV 2 infection.

Immune response due to malaria is bimodal in nature, which includes Th1-type response for limiting initial parasitemia and further Th2 based cytokine production for parasite clearance. It is a common assumption that immunity against malaria parasite is short-lived and that a continuous exposure to parasite antigens is needed to keep it alive. However, several studies came up which differ from such observations and have pointed out that in endemic areas anti-malarial immunity survive for longer durations [17] [18] [19] [20] [21] [22] [23]. It has been observed that immigrants from endemic areas maintain some clinical immunity against clinical malaria [24] [25] [26]. It is also been observed in epidemiological studies that in areas of low and unstable malaria transmission, such as South Africa, individuals having prior exposures even decades ago, can have protective immunity much later in life [27].

Reemergence of malaria in Madagascar after a long period of total control, was not able to infect people present 30 years earlier during malaria exposure. These Individuals were much more resistant to clinical disease than were younger population, which points to an aspect of immunity which is long lived [23].

Studies in malaria endemic regions has pointed out towards sustained levels of IFN-γ and IL-10 cytokines upto 3 years and 6 years respectively [28] [29]. In experimental malaria, single *P. falciparum* infection can approach the persistent of IFN-γ and γδ T cells which remain viable up to 14 months [27] [30], whereas under natural infections the half-life for the IFN-γ and yδ T cells is around 3.3 years [31], with IL-10 and CD4^+^ T cells maintained till 6 years [31]. Further, T_reg_ cells in acute malaria episodes are also been suggested to expanded from pre-exsisting pool of memory T cells [32]. As the analysis in our study pointed out about less prevalence of COVID-19 infection in malaria endemic regions of the globe, it will be relevant to consider role of immune-response against malaria in providing resistance to the infection.

One may argue that the high malaria endemic regions are relatively warmer than the regions worst affected by COVID-19 pandemic, but from overall consensus from past and current data, it appears that SARS-CoV-2 can be transmitted in hot and humid weather as well [33].

In view of above mentioned literature we hypothesize that a pre-existing immunity, due to malaria infection, in the malaria endemic countries is responsible for slow growth of SARS-CoV-2 infections. However, the part of globe which has never faced malaria transmission have not shown such resistance to the SARS-CoV-2 infection and hence the high mortality and infections are been observed.

## Data Availability

We have used World Malaria Report 2019 by WHO on malaria and Worldometer data on COVID-19.

## Notes

### Competing Interest Statement

The authors have declared no competing interest.

### Funding Statement

We are thankful to ICMR and DBT GoI for providing funding.

## References

1. Khosrawipour, V., et al., Failure in initial stage containment of global COVID-19 epicenters. J Med Virol, 2020.

2. Maire, N., et al., A model for natural immunity to asexual blood stages of Plasmodium falciparum malaria in endemic areas. Am J Trop Med Hyg, 2006. 75(2 Suppl): p. 19–31.

3. Snow, R.W., et al., The global distribution of clinical episodes of Plasmodium falciparum malaria. Nature, 2005. 434(7030): p. 214–7.

4. Doolan, D.L., C. Dobano, and J.K. Baird, Acquired immunity to malaria. Clin Microbiol Rev, 2009. 22(1): p. 13–36, Table of Contents.

5. Moorlag, S., et al., Non-specific effects of BCG vaccine on viral infections. Clin Microbiol Infect, 2019. 25(12): p. 1473–1478.

6. Bustinduy, A.L., et al., Age-Stratified Profiles of Serum IL-6, IL-10, and TNF-alpha Cytokines Among Kenyan Children with Schistosoma haematobium, Plasmodium falciparum, and Other Chronic Parasitic Co-Infections. Am J Trop Med Hyg, 2015. 92(5): p. 945–51.

7. Schofield, L. and I. Mueller, Clinical immunity to malaria. Curr Mol Med, 2006. 6(2): p. 205–21.

8. Chaves, Y.O., et al., Immune response pattern in recurrent Plasmodium vivax malaria. Malar J, 2016. 15(1): p. 445.

9. Shen, S.S., et al., Infection against infection: parasite antagonism against parasites, viruses and bacteria. Infect Dis Poverty, 2019. 8(1): p. 49.

10. Bilenko, N., et al., Does co-infection with Giardia lamblia modulate the clinical characteristics of enteric infections in young children? Eur J Epidemiol, 2004. 19(9): p. 877–83.

11. Teo, T.H., et al., Plasmodium co-infection protects against chikungunya virus-induced pathologies. Nat Commun, 2018. 9(1): p. 3905.

12. Frieman, M., M. Heise, and R. Baric, SARS coronavirus and innate immunity. Virus Res, 2008. 133(1): p. 101–12.

13. Cameron, M.J., et al., Interferon-mediated immunopathological events are associated with atypical innate and adaptive immune responses in patients with severe acute respiratory syndrome. J Virol, 2007. 81(16): p. 8692–706.

14. Zhang, Y., et al., Analysis of serum cytokines in patients with severe acute respiratory syndrome. Infect Immun, 2004. 72(8): p. 4410–5.

15. Jiang, Y., et al., Characterization of cytokine/chemokine profiles of severe acute respiratory syndrome. Am J Respir Crit Care Med, 2005. 171(8): p. 850–7.

16. Yu, S.Y., et al., Gene expression profiles in peripheral blood mononuclear cells of SARS patients. World J Gastroenterol, 2005. 11(32): p. 5037–43.

17. Matteelli, A., et al., Epidemiological features and case management practices of imported malaria in northern Italy 1991-1995. Trop Med Int Health, 1999. 4(10): p. 653–7.

18. Jelinek, T., et al., Imported Falciparum malaria in Europe: sentinel surveillance data from the European network on surveillance of imported infectious diseases. Clin Infect Dis, 2002. 34(5): p. 572–6.

19. MacMullin, G., et al., Host immune response in returning travellers infected with malaria. Malar J, 2012. 11: p. 148.

20. Mascarello, M., et al., Imported malaria in adults and children: epidemiological and clinical characteristics of 380 consecutive cases observed in Verona, Italy. J Travel Med, 2008. 15(4): p. 229–36.

21. Bouchaud, O., et al., Do African immigrants living in France have long-term malarial immunity? Am J Trop Med Hyg, 2005. 72(1): p. 21–5.

22. Salvado, E., et al., [Clinical presentation and complications of Plasmodium falciparum malaria in two populations: travelers and immigrants]. Enferm Infecc Microbiol Clin, 2008. 26(5): p. 282–4.

23. Monge-Maillo, B., et al., Plasmodium falciparum in asymptomatic immigrants from sub-Saharan Africa, Spain. Emerg Infect Dis, 2012. 18(2): p. 356–7.

24. Kleinschmidt, I., et al., Use of generalized linear mixed models in the spatial analysis of small-area malaria incidence rates in Kwazulu Natal, South Africa. Am J Epidemiol, 2001. 153(12): p. 1213–21.

25. Luxemburger, C., et al., The epidemiology of severe malaria in an area of low transmission in Thailand. Trans R Soc Trop Med Hyg, 1997. 91(3): p. 256–62.

26. Deloron, P. and C. Chougnet, Is immunity to malaria really short-lived? Parasitol Today, 1992. 8(11): p. 375–8.

27. Teirlinck, A.C., et al., Longevity and composition of cellular immune responses following experimental Plasmodium falciparum malaria infection in humans. PLoS Pathog, 2011. 7(12): p. e1002389.

28. Moormann, A.M., et al., Stability of interferon-gamma and interleukin-10 responses to Plasmodium falciparum liver stage antigen 1 and thrombospondin-related adhesive protein immunodominant epitopes in a highland population from Western Kenya. Am J Trop Med Hyg, 2009. 81(3): p. 489–95.

29. Moormann, A.M., et al., Stability of interferon-gamma and interleukin-10 responses to Plasmodium falciparum liver stage antigen-1 and thrombospondin-related adhesive protein in residents of a malaria holoendemic area. Am J Trop Med Hyg, 2006. 74(4): p. 585–90.

30. Sauerwein, R.W., M. Roestenberg, and V.S. Moorthy, Experimental human challenge infections can accelerate clinical malaria vaccine development. Nat Rev Immunol, 2011. 11(1): p. 57–64.

31. Wipasa, J., et al., Short-lived IFN-gamma effector responses, but long-lived IL-10 memory responses, to malaria in an area of low malaria endemicity. PLoS Pathog, 2011. 7(2): p. e1001281.

32. Walther, M., et al., Distinct roles for FOXP3 and FOXP3 CD4 T cells in regulating cellular immunity to uncomplicated and severe Plasmodium falciparum malaria. PLoS Pathog, 2009. 5(4): p. e1000364.

33. Luo, W., et al., The role of absolute humidity on transmission rates of the COVID-19 outbreak.medRxiv 17 feburary 2020. doi:https://doi.org/10.1101/2020.02.12.20022467

